# Evaluating the impact of modeling choices on the performance of integrated genetic and clinical models

**DOI:** 10.1101/2023.11.01.23297927

**Authors:** Theodore J. Morley, Drew Willimitis, Michael Ripperger, Hyunjoon Lee, Lide Han, Yu Zhou, Jooeun Kang, Lea K. Davis, Jordan W. Smoller, Karmel W. Choi, Colin G. Walsh, Douglas M. Ruderfer

## Abstract

The value of genetic information for improving the performance of clinical risk prediction models has yielded variable conclusions. Many methodological decisions have the potential to contribute to differential results across studies. Here, we performed multiple modeling experiments integrating clinical and demographic data from electronic health records (EHR) and genetic data to understand which decision points may affect performance. Clinical data in the form of structured diagnostic codes, medications, procedural codes, and demographics were extracted from two large independent health systems and polygenic risk scores (PRS) were generated across all patients with genetic data in the corresponding biobanks. Crohn’s disease was used as the model phenotype based on its substantial genetic component, established EHR-based definition, and sufficient prevalence for model training and testing. We investigated the impact of PRS integration method, as well as choices regarding training sample, model complexity, and performance metrics. Overall, our results show that including PRS resulted in higher performance by some metrics but the gain in performance was only robust when combined with demographic data alone. Improvements were inconsistent or negligible after including additional clinical information. The impact of genetic information on performance also varied by PRS integration method, with a small improvement in some cases from combining PRS with the output of a clinical model (late-fusion) compared to its inclusion an additional feature (early-fusion). The effects of other modeling decisions varied between institutions though performance increased with more compute-intensive models such as random forest. This work highlights the importance of considering methodological decision points in interpreting the impact on prediction performance when including PRS information in clinical models.

## Introduction

Multiple recent studies have sought to integrate genetic information, often in the form of polygenic risk scores (PRS), which index the genome-wide contributions of common variants, with clinical data to establish models that improve risk stratification and clinical prediction^1–13^. These studies hypothesize that genetic information may enhance prediction of clinical outcomes above and beyond the performance of models using only clinical features. Further, since a patient’s genome can be assayed at birth, this information provides a potential opportunity to understand risk and intervene prior to the development of disease. However, the results of these efforts have offered conflicting evidence on the value of PRS, with some supporting the benefits of including genetic information and others finding no benefit at all.

The focus on whether genetic information improves clinical risk prediction obscures the many methodological decisions that might impact these results. In addition to the properties of the selected phenotype (e.g. prevalence, severity), prediction models depend on numerous decisions that could impact performance. These include 1) whether genetic data are incorporated as additional feature(s) at the time of model training (“early fusion^14^”) or combined with the predictions from an existing non-genetic model (“late fusion^14^”), 2) whether to use a model trained in a larger, broader patient population or within the specific targeted population, 3) which statistical methods are used for model building from pre-processing to model selection and tuning, and 4) how model performance is quantified and evaluated.

The increasing accessibility and use of electronic health records (EHR) have provided an opportunity to leverage real world clinical data to establish prediction models to inform risk^15^. The linking of biobanks to these EHRs enables the inclusion of genetic information at scale for assessment of models using both clinical and genetic data. Although incorporating genomic data requires pragmatic considerations (e.g., cost and time to acquire, selection bias, etc.,), this information can be input alongside other known risk factors to generate prediction models. Importantly, studies are already prospectively collecting genetic data and providing results back to patients clinically to assess their utility^16,17,18^.

Here, we sought to assess the impact of methodological decisions on predictive performance when combining clinical data from EHR with genetic information in the form of polygenic risk scores. An ideal phenotype to test the impact of these decisions would have substantial genetic contribution, be sufficiently prevalent^19^, and readily defined from high quality phenotypic definitions^20^ derived from EHRs. One such phenotype is Crohn’s disease, a heritable inflammatory bowel disease that is common (age standardized prevalence = 0.42%)^21^ and for which EHR-based phenotyping algorithms have been established^22^. Rigorously evaluating these modeling decisions might inform strategies to develop integrated models of clinical and genetic data for predicting a wide range of phenotypes while also providing better understanding of the impact of genetic information in predicting the onset of Crohn’s Disease.

## Methods

### Study population

Clinical and genetic data were extracted from the EHRs and biobanks of Vanderbilt University Medical Center (VUMC) and Mass General Brigham (MGB) as part of the PsycheMERGE^23^ network. VUMC is an academic medical center in Nashville, Tennessee, that manages over 2 million patient visits every year across Tennessee and its neighboring states. The Synthetic Derivative (SD)^11^ is a deidentified repository used to store clinical EHR data at VUMC. The VUMC biobank (BioVU) is directly linked to the SD and includes over 300,000 DNA samples. MGB is a major healthcare system including Massachusetts General Hospital, Brigham and Women’s Hospital, and other community and specialty hospitals located in Boston, Massachusetts. The deidentified clinical EHR data were extracted from the MGB Research Patient Data Registry (RPDR)^24^, a data warehouse that covers over 20 years of data from more than 6.5 million patients. The MGB biobank (MGBB) is linked to RPDR and includes over 80,000 DNA samples. For each study site, we defined two independent samples: 1) those with only clinical information from the EHR but no available genetic information (“EHR only”) and 2) those with both EHR and genetic information (“genetic sample”). All analyses were approved by the institutional review boards at each site.

### Phenotype definition

We defined cases at each site based on a phenotyping algorithm for Crohn’s disease from PheKB^25^ excluding the use of clinical notes. Specifically, cases were patients who had at least two independent ICD-9/10 codes for Crohn’s disease and at least one of a list of medications used to treat Crohn’s disease, e.g., methotrexate. Because other gastrointestinal or autoimmune diseases may occasionally be misdiagnosed as Crohn’s disease, we excluded individuals who had any diagnoses of ulcerative colitis or other autoimmune conditions. Controls included all patients who had never received any Crohn’s disease diagnostic code as defined above. A minimum data criterion was applied at each site requiring patients to have at least 2 visits over 30 days apart before the censoring date (24 hours before the first Crohn’s disease diagnosis date for cases and the last diagnosis date for controls).

### Demographic and clinical features extracted from EHRs

Candidate predictors at VUMC were drawn from routinely collected EHR data including: demographics (age at censoring date in years, categorical coded sex [male, female, unknown], coded race [White, Black, Asian, Other, Unknown]), coded ethnicity [Hispanic, Not-Hispanic, Unknown], diagnostic codes (log-transformed counts of historical Clinical Classification Software Level 2 codes^26^), medications (log-transformed counts of RXNORM-mapped ingredients^27^) and body mass index. Candidate predictors at MGB were the same as those of VUMC except for BMI which was not available. Missing BMI values were imputed using a single multivariable imputation by the aregImpute function from the Hmisc package^28^. Diagnostic codes and medications given after the first Crohn’s disease diagnosis code for cases and last diagnosis code for controls were removed.

### Polygenic risk score (PRS) calculation

Crohn’s disease PRS were calculated in genotyped patients from both VUMC and MGB using the summary statistics from the most recent IBD GWAS^29^. PRS scoring was performed using PRS-CS which places a continuous shrinkage prior on SNP effect sizes using a Bayesian regression framework^30^. The continuous shrinkage priors adapt the amount of shrinkage applied to each SNP to the strength of the associated GWAS signal based on the LD structure estimated from an external reference panel. After ancestry estimation from principal component analysis, sample sizes were large enough only in the European ancestry sample. The 1,000 Genomes European reference panel was used to estimate LD between SNPs. The PRS were summed for each individual of the target cohort using Plink 1.9^31,32^. A linear regression was performed to derive a residualized PRS after including the first 10 principal components of genetic ancestry.

### EHR-based prediction models

Models were trained separately at both sites using either the EHR-only sample or the genetics sample, but all models were evaluated on the genetics sample. In total, 10 models were trained with each addressing a specific scientific question (**Supplementary Table 1**). The first three models tested the baseline performance of PRS, demographics and the combination of the two. Specifically, a logistic regression using the Crohn’s PRS (Model 1) was used to assess performance of PRS alone, a random forest classifier was trained using only demographics (age at censoring date, coded sex, BMI, coded race, and coded ethnicity; Model 2) and a logistic regression of the demographic data and the PRS (Model 3) was used to test the value of PRS when added to the demographic information. Next, to test the impact of training sample on performance we generated two L1-penalized regression models using all demographic and clinical features trained on the EHR-only sample (Model 4) or trained on the genetics sample (Model 5). We then developed two late-fusion models to assess the impact of PRS on each of these clinical models simply by including the PRS in a logistic regression including an interaction term between the two features based on the EHR-only sample (Model 6) and the genetics sample (Model 7). Our final 3 models leveraged more compute-intensive classifier selection approaches which evaluated multiple different classification methods to generate a clinical only model (Model 8) and to compare two different methods on integrating the genetic information: late-fusion (Model 9) and early-fusion (Model 10). Model 8 was a random forest selected classifier using all demographic and clinical features trained on the genetic sample. Model 9 used a logistic regression approach to integrate the clinical prediction score from Model 8 with the PRS and Model 10 was also a random forest classifier that included the PRS as an additional feature in model training.

For models trained on the EHR-only sample, we refit the model and generated out of sample predictions on the independent genetic sample. For models trained on the genetic sample we applied 5-fold nested-cross validation as follows. We first used an “outer” stratified cross-validation loop to produce “outer” train and “outer” test folds. Then, within each iteration of this “outer” loop, we applied an “inner” cross-validation with randomized hyperparameter search on the “outer” training fold and used the best model parameters to predict on the “outer” test fold for each classification algorithm^33^. After this process is finished, out of sample predictions are produced from each optimized classification algorithm for the entire genetic sample resulting in five sets of predictions and performance metrics. Models 2, 3, 8, 9, and 10 further implemented a model development and selection routine that evaluated three additional classifier types – logistic regression, random forest, and adaboost. Scikit-Learn^34^ was used for all modeling procedures.

In order to meet assumptions for logistic regression when used in late fusion we applied a quantile transformation to generate a normal distribution on the predicted probabilities from the initial clinical model, and used the standard scaler from scikit-learn^34^ on the PRS. In the clinical model, we also used the standard scaler on BMI and age. We found that the highest performance was gained when including these preprocessing steps.

Model performance was evaluated with discrimination metrics: Area Under the Receiver Operating Characteristic (AUROC) and Area Under the Precision-Recall Curve (AUPRC). To evaluate performance of the methods overall, we calculated the mean across sets for both AUROC and AUPRC.

### Quantifying impact of PRS on performance overall and stratified by age

To assess the change in model performance due to inclusion of the PRS, we calculated the Integrated Discrimination Index (IDI)^35^, and the Net Reclassification Improvement (NRI)^35^ which both aim to quantify the impact of new information on a predictive method, with the IDI doing so through the shift in mean probabilities and the NRI doing so through examining the proportions of both cases and controls whose probabilities shift either upwards or downwards. Both methods are sensitive to differences in calibration as a product of the different classification methods. To correct for these differences, we performed logistic calibration^36^ to recalibrate the probabilities and more accurately compare with the late fusion regression model.

To assess whether PRS was having a disproportionate impact among demographic groups (age and sex), we additionally stratified the cohort by sex and below or above the median age of first Crohn’s diagnosis at VUMC (30 years old) and MGB (47 years old) and calculated ΔAUPRC between models, which we calculated by taking the AUPRC for both the clinical only model and the clinical+PRS model in each scenario and finding the difference between them to evaluate the improvement from adding PRS.

## Results

### Comparison of baseline genetic and demographic models for Crohn’s disease across two institutions

Patients with Crohn’s disease were identified from EHR data based on presence of diagnostic codes and medications specific to Crohn’s and exclusive of ulcerative colitis (see Methods). Across two healthcare institutions, VUMC and MGB, 4,786 patients met algorithm criteria for Crohn’s disease (VUMC: 2,366, MGB: 2,566). Patients were subsequently divided into those with corresponding genetic data (“genetic sample”) and those without (“EHR only”). Among the samples with genetic information, we observed 189 cases at VUMC and 53 cases at MGB, representing outcome prevalence rates of 0.35% and 0.22%, respectively. Controls in the EHR only sample were randomly selected to match the prevalence rates in the genetics sample (VUMC: 53,496, MGB: 23,669, **Table 1**).

**Table 1:** Demographics of the EHR and genetic samples at VUMC and MGB. BMI data were not available at MGB.

|  |  | *Full EHR* |  |  |  | *Genetic sample* |  |  |  |
| --- | --- | --- | --- | --- | --- | --- | --- | --- | --- |
|  |  | VUMC |  | MGB |  | VUMC |  | MGB |  |
| Demographic |  | Crohn's Disease (N=1,072) | Control (N=559,416) | Crohn's Disease (N=2,566) | Controls (N=1,145,936) | Crohn's Disease (N=189) | Control (N=53,495) | Crohn's Disease (N=53) | Controls (N=23,669) |
| Age | Mean at index time | 34.83 | 43.28 | 44.25 | 48.63 | 33.28 | 53.89 | 48.43 | 62.3 |
| Sex | Female | 547 | 309,925 | 1,345 | 647,635 | 96 | 29,991 | 29 | 12,553 |
|  | Male | 525 | 249,477 | 1,220 | 498,241 | 93 | 23,504 | 24 | 11,116 |
|  | Unknown | 0 | 14 | 1 | 60 | 0 | 0 | 0 | 0 |
| Race | White | 898 | 424,067 | 2,224 | 859,127 | 188 | 50831 | 53 | 23,145 |
|  | Black | 122 | 64,043 | 89 | 73,670 | 0 | 42 | 0 | 7 |
|  | Asian | 15 | 9,808 | 59 | 54,169 | 0 | 20 | 0 | 4 |
|  | Other | 37 | 61,498 | 194 | 158,970 | 1 | 2602 | 0 | 513 |
| Ethnicity | Hispanic | 20 | 20,609 | 42 | 52,739 | 0 | 246 | 0 | 2 |
|  | Non-Hispanic | 1,028 | 484,257 | 2,524 | 1,093,197 | 186 | 50846 | 53 | 23667 |
|  | Unknown | 24 | 54,550 | NA | NA | 3 | 2403 | 0 | 0 |
| BMI | Mean | 28.65 | 27.45 | NA | NA | 26.35 | 28.51 | NA | NA |

To establish baseline comparisons, we evaluated the independent predictive performance of a model including only the Crohn’s disease PRS (Model 1, **Table 2**) or only demographic features (Model 2). The PRS only model yielded an AUROC of 0.68 for VUMC and 0.67 for MGB and AUPRC of 0.018 and 0.011 for VUMC and MGB, respectively (**Figure 1**). The model with demographic information alone had AUROC of 0.75 for VUMC and 0.74 for MGB and AUPRC of 0.013 and 0.028 at VUMC and MGB, respectively. A model that included PRS and demographic features (Model 3), achieved higher AUROCs (VUMC: 0.80, MGB: 0.76) and AUPRCs (VUMC: 0.029, MGB: 0.046) at both sites.

**Figure 1.**
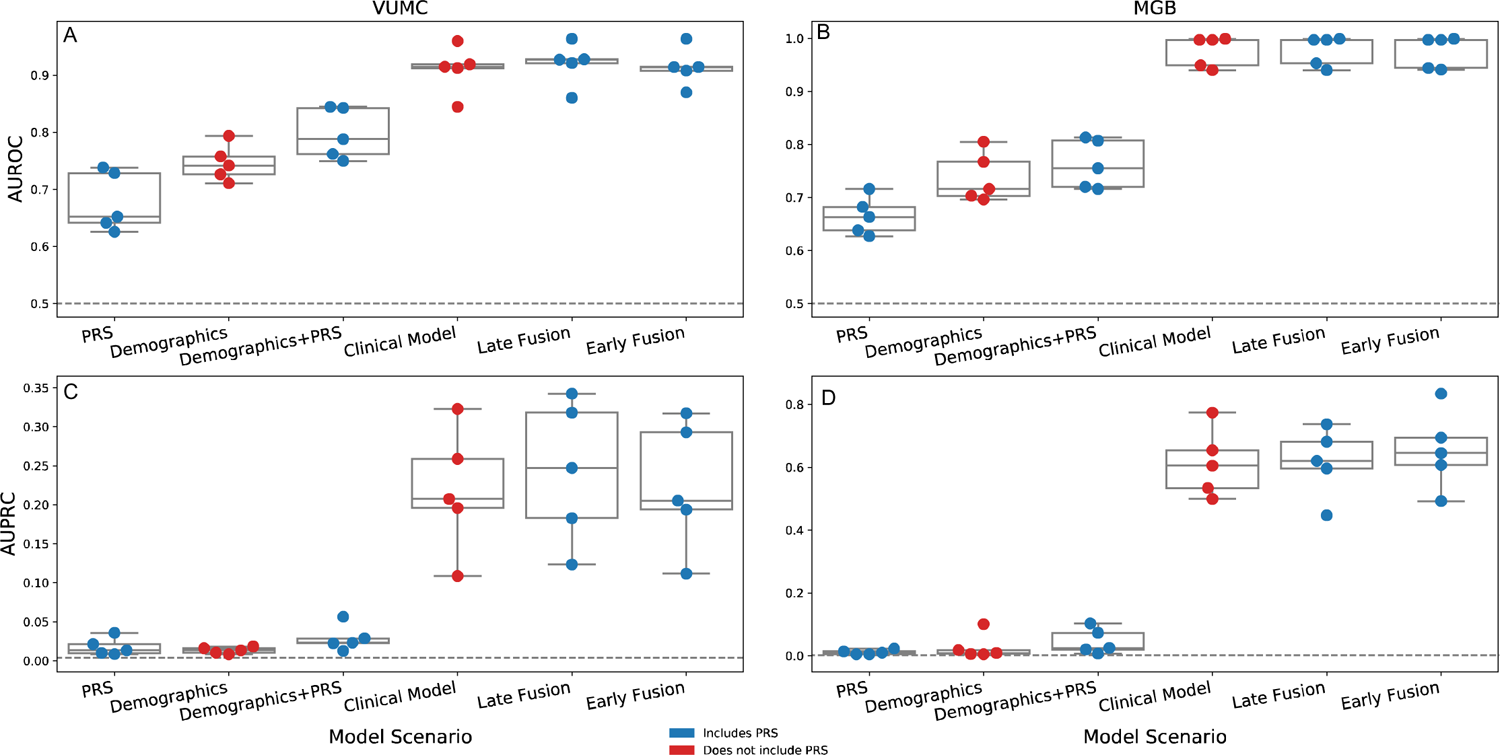
Performance metrics (top: AUROC, bottom: AUPRC) are presented within each institution (left: VUMC, right: MGB) for each model scenario. Colors represent whether the model included PRS (Blue) or was based only on EHR data (Red). Models run on the genetics sample required five-fold nested cross-validation and each point represents the on split that is consistent across all models for comparability. Dashed lines represent the baseline performance in each site for the given metric.

**Table 2:** AUROC and AUPRC at VUMC and MGB, for each of the primary models. Each modelling approach was evaluated across five splits in the nested cross validation approach, and performance is reported by the mean value and the range across the splits.

| Model Names |  | VUMC |  |  |  | MGB |  |  |  |
| --- | --- | --- | --- | --- | --- | --- | --- | --- | --- |
|  |  | AUROC |  | AUPRC |  | AUROC |  | AUPRC |  |
|  |  | Mean | Range | Mean | Range | Mean | Range | Mean | Range |
| Model 1 | PRS | 0.68 | 0.63-0.74 | 0.018 | 0.0084-0.036 | 0.67 | 0.63-0.72 | 0.011 | 0.005-0.023 |
| Model 2 | Demographics | 0.75 | 0.71-0.80 | 0.013 | 0.0083-0.018 | 0.74 | 0.70-0.81 | 0.028 | 0.005-0.10 |
| Model 3 | PRS+Demo | 0.80 | 0.75-0.84 | 0.029 | 0.013-0.087 | 0.76 | 0.72-0.81 | 0.046 | 0.007-0.10 |
| Model 8 | Clinical Model | 0.91 | 0.84-0.96 | 0.22 | 0.11-0.32 | 0.98 | 0.94-0.99 | 0.61 | 0.50-0.77 |
| Model 9 | Late Fusion | 0.92 | 0.86-0.96 | 0.24 | 0.12-0.34 | 0.98 | 0.94-0.99 | 0.62 | 0.44-0.74 |
| Model 10 | Early Fusion | 0.91 | 0.87-0.96 | 0.22 | 0.11-0.29 | 0.98 | 0.94-0.99 | 0.64 | 0.49-0.83 |

### Minimal increase in performance seen when adding PRS to clinical models

We incorporated the PRS into our best-performing clinical prediction model using two approaches: 1) a “late-fusion” approach where a model is trained on only the PRS and the predicted probability of the clinical model, and 2) an “early-fusion” approach in which the model is trained with all clinical features plus the PRS. The best performing clinical model (Model 8) performed well at both VUMC (AUROC = 0.91, AUPRC = 0.22) and MGB (AUROC = 0.98, AUPRC = 0.61). Integrating the PRS with that model using late-fusion (Model 9) resulted in improved average performance in both VUMC (AUROC = 0.92, AUPRC=0.24) and MGB (AUROC = 0.98, AUPRC=0.62). The best performing early fusion model at VUMC provided the same performance as the clinical model alone (AUROC = 0.91, AUPRC = 0.22). At MGB, the early fusion model had the same average AUROC but higher average AUPRC than the late fusion model (AUROC=0.98, AUPRC=0.64).

### PRS improves discrimination over demographic features alone but not in models with high-dimensional clinical information

To quantitatively assess the impact of adding PRS, we calculated the change in performance metrics (ΔAUROC, ΔAUPRC) as well as the integrated discrimination index (IDI) and net reclassification index (NRI) for both late and early fusion approaches (Model 9 and 10) compared to the clinical model that they are fused with (Model 8). As a comparator, we included the scenario where PRS was integrated with the demographics only (Model 3, Figure 2). Adding PRS to the demographics only model resulted in consistent improvements across splits for all metrics at both VUMC (ΔAUROC = 0.051, ΔAUPRC = 0.015, NRI = 0.58, IDI = 0.0054) and MGB (ΔAUROC = 0.025, ΔAUPRC = 0.018, NRI = 0.45, IDI = 0.0026). For late-fusion, consistent positive values were seen for ΔAUROC and NRI but IDI and ΔAUPRC spanned zero in VUMC and all metrics spanned zero for MGB (Figure 2). No metric showed consistent improvement for early-fusion at either VUMC or MGB.

**Figure 2.**
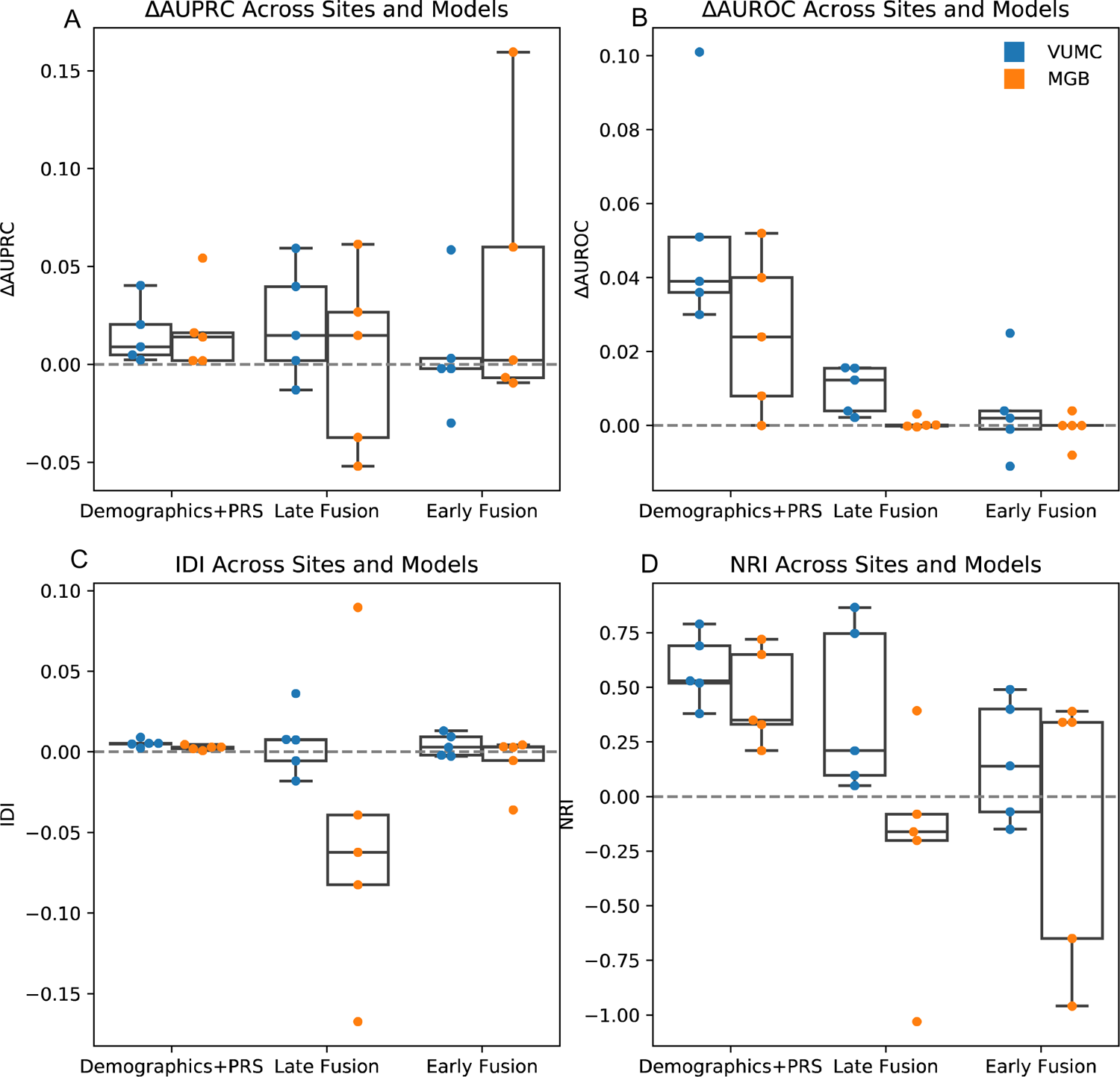
Change within ΔAUPRC (A), ΔAUROC (B), IDI (C), and NRI (D) after the integration of genetic information in each scenario, and within each site.

### PRS has most value among young Crohn’s patients in model combined with demographics only

We examined changes in AUPRC stratified by coded sex and age (above vs. below median age at first diagnosis, 30 at VUMC, 47 at MGB) to assess whether PRS had a stronger effect on performance in a particular subgroup of the sample when only including demographic information (Figure 3). A consistent increase in performance was seen among the younger half of the sample in both VUMC and MGB (Figure 3). No difference was observed by sex.

**Figure 3:**
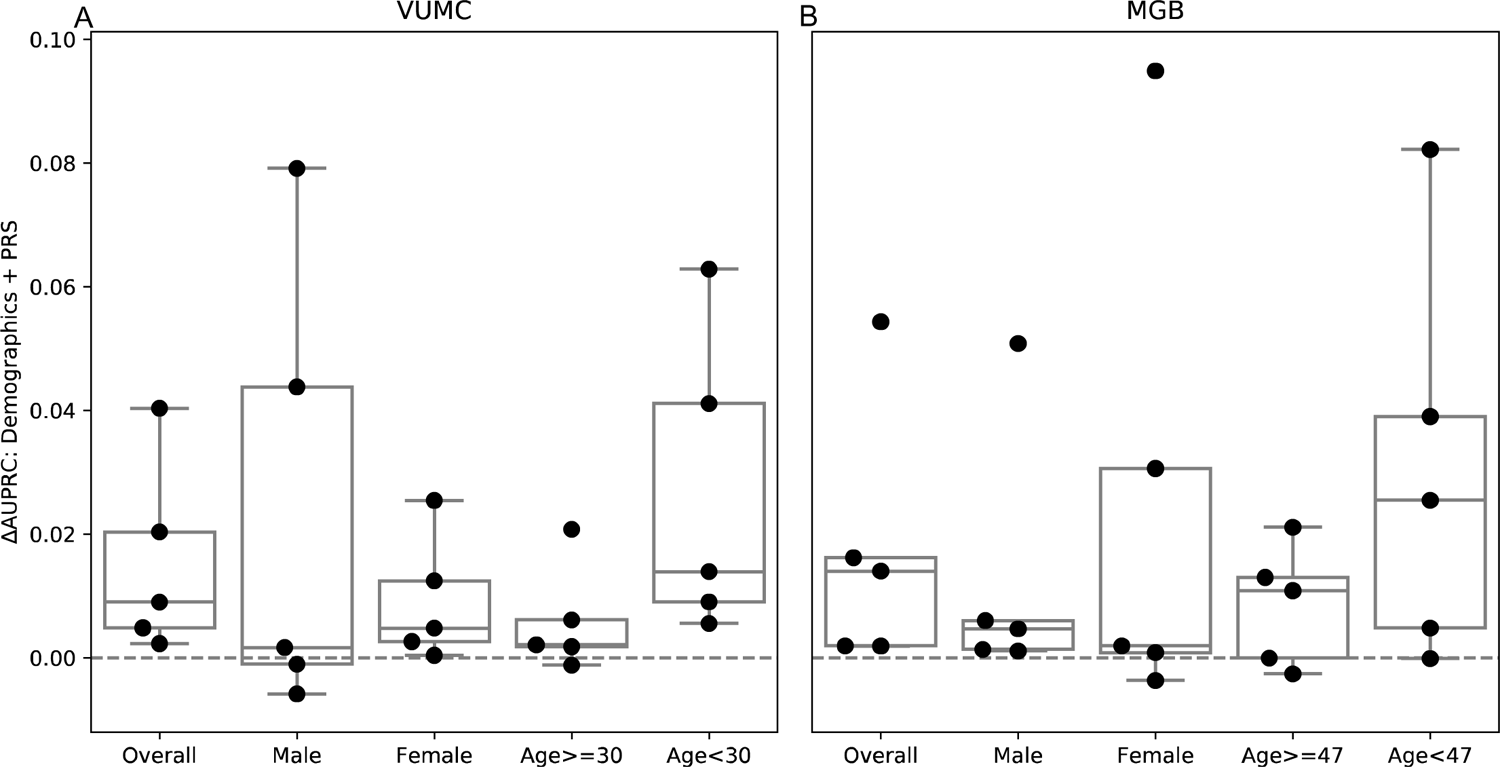
Change in performance for each subgroup at VUMC (A) and MGB (B) when adding genetic information to the demographic information only model (includes BMI at VUMC).

## Discussion

The increasing availability of genetic information presents a potential avenue to improve clinical risk prediction. However, published evidence to date on the benefit of genetic information on predictive performance has been inconsistent. Here, we aimed to better understand the implications of different methodological decisions in integrating clinical data from the EHR with genetic data for risk prediction using the example of Crohn’s disease. We examined the effect of varying training sample (see supplement), classifier selection, performance metric and method of combining PRS with clinical variables on model performance. We observed consistent performance gains when introducing more complex classification methods (e,g. using random forest vs. logistic regression) and from adding PRS to only demographic information. The inclusion of clinical information substantially reduces the incremental predictive value of the PRS, with late-fusion outperforming early-fusion but only in a subset of performance metrics at one of the sites.

Improvement in predictive performance and discrimination is clear across all metrics when adding PRS to demographic information alone. The greatest marginal benefit of adding PRS to demographic features was seen among younger patients (i.e., those below the median of age), consistent with the expected relationship of greater genetic influence among those with earlier onset of disease. This finding suggests the incremental value of PRS might be greater when limited longitudinal clinical data are available or when existing clinical risk models have relatively poor performance (such that any improvement in predictive performance might be useful). Indeed, the value of genetic information largely disappears when the clinical information is included, regardless of approach to integration. While the clinical data may reflect expression of underlying genetic risk, once disease manifests sufficient to be reflected in clinical data, genetic information provides less value. Identifying the optimal clinical timepoints where incorporating genetic information (e.g., when a patient is first seen in the system, or in early life) would be add value remains an important open question.

To compare different integration strategies, we built models using both late and early fusion of PRS with clinical data. Early-fusion did not show significant performance improvement across any metric at either site. In the presence of high-dimensional clinical features, much of which is likely correlated with genetic risk, it may be that the addition of a single PRS feature provides little additional signal. Late-fusion also showed minimal benefit, though with nominal improvement in AUROC and IDI at one site (VUMC). Although neither integration approach produced meaningful performance improvements in this study, the finding that late-fusion performed as well or better than early fusion is notable as late-fusion (which adds PRS to existing model predictions) provides a more time- and cost-effective approach for combining PRS and clinical information. For example, the full clinical model at VUMC took approximately 32 hours in training and cross-validation as well as a significant amount of development work to set up compared to the late-fusion model which can be run in minutes.

We chose to include four metrics that capture different aspects of predictive performance and discrimination. Our results illustrate that the interpretation of incremental value of incorporating PRS in clinical models may depend on how performance is evaluated. When all four metrics consistently show improvement as in the case of adding PRS to demographics, it is easy to interpret that PRS is improving performance. In other situations, such as late-fusion, that interpretation might depend on which specific metric was used with potential value being seen at VUMC from ΔAUROC and NRI but not ΔAUPRC or IDI. The over reliance on any single metric or the over interpretation of any numerical increase might not be robust. Ultimately, specific analyses that quantify the improvement in clinical utility as a product of including genetic information will be required.

There are several limitations to this work. First, our methodologic comparisons focused on a single exemplar phenotype, Crohn’s disease, chosen because of its established polygenic basis and the availability of a validated algorithm for EHR-based phenotyping. Whether our findings apply to risk models for other diseases remains to be established. Second, the precision of our performance metrics was constrained by available sample sizes for the test sets despite our examination of two large healthcare systems and their biobanks. Third, reflecting the limited ancestral diversity of the participating biobanks, our analyses were restricted to patients of European ancestry, and it will be important for future studies to evaluate the applicability of our results in transancestry prediction models. There are several barriers to the immediate uptake of genetics in the clinical beyond utility including but not limited to cost and known disparities in prediction performance among different ancestral groups^37^.

This work provides an example of the implications of important methodological decision making when designing and implementing a clinical prediction model incorporating genetic information. Overall, we find that incorporating PRS into clinical prediction models, whether by early- or late-fusion, provides limited improvements in performance. Future work is needed to determine under which circumstances and for which diseases incorporating PRS may have the biggest impact. Ultimately, incorporating genetic information in clinical prediction models will likely become more common and the impact of decision points described here can inform the design and implementation of those models.

## Data Availability

All data produced in the present work are contained in the manuscript. Sharing of individual level data is restricted.

## Acknowledgements

This work was supported in part by grant 1R01MH118233 from the National Institutes of Mental Health (NIMH). The dataset(s) used for the analyses described were obtained from Vanderbilt University Medical Center’s BioVU, which is supported by institutional funding, private agencies, and federal grants. These include NIH funded Shared Instrumentation Grant S10OD017985, S10RR025141, and S10OD025092; CTSA grants UL1TR002243, UL1TR000445, and UL1RR024975. Genomic data are also supported by investigator-led projects that include U01HG004798, R01NS032830, RC2GM092618, P50GM115305, U01HG006378, U19HL065962, R01HD074711; and additional funding sources listed at https://victr.vanderbilt.edu/pub/biovu/. We thank Mass General Brigham Biobank for providing samples, genomic data, and health information data. This work was conducted in part using the resources of the Advanced Computing Center for Research and Education at Vanderbilt University, Nashville, TN.

