## Supplementary Methods for "Evaluating the impact of modeling choices on the performance of integrated genetic and clinical models"

**Supplementary Information**

**Supplementary Methods**

**Establishing models using different training samples (EHR only vs genetic)**

We further considered the potential impact of the source of model training data, using our EHR only sample to represent an external dataset, and our genetic sample as our internal dataset. By EHR only sample, we mean the group of individuals within our hospital system who have no available genetic data for us to use, but who do have clinical data such as diagnostic codes and medications available. The genetic sample, comparatively, has access to the same clinical data, but additionally has genetic information available. We did not see large differences in performance across institutions (Supplementary Table 1).

**Supplementary Results**

**Comparison of models trained on the EHR only sample versus genetics sample**

In addition to the model using a full classifier selection process, we also examined the performance of late-fusion models when only using penalized regression (LASSO), where the clinical model was trained either in the EHR-only sample or the genetics sample, with the goal of comparing whether there were benefits depending on the training source, such as the increased amount of data from the EHR-sample, or if the misspecification from training on patients selected differently (EHR-only) would negatively impact the outcome. In general, we saw slightly better performance in the genetics sample than the EHR sample at VUMC, and slightly worse performance in the genetics sample than in the EHR sample at MGB (Supplementary Table 1).
